# Insomnia and the genetic overlap of sleep with depression and anxiety: a genomic structural equation modelling study

**DOI:** 10.64898/2026.10.21.26366134

**Authors:** Mehmet Akif Şakiroğlu, Yusuf Çiçek, Murat Can Çelik, Ali Tarık Altunç, Doğukan Aslan, Halil Aziz Velioğlu

## Abstract

Insomnia, depression and anxiety co-occur and share genetic liability, but a genetic correlation cannot show whether that liability is spread across the sleep domain or concentrated in insomnia, a diagnostic criterion for depression and generalized anxiety disorder. We applied genomic structural equation modelling to ten genome-wide association studies and fitted a four-factor model of sleep, internalizing (depression and anxiety), psychotic-bipolar and metabolic liability. A model linking the sleep and internalizing domains only through their factor correlation was inadmissible, and we compared, under one decision rule, each one-parameter link that could repair it: domain-wide links, an insomnia-specific cross-loading, and residual covariances representing shared diagnostic content, of which three were admissible. The insomnia cross-loading fitted better than the only admissible domain-wide alternative (ΔAIC = 13.8; better in 98.6% of 500 Monte Carlo draws) and than shared content (ΔAIC = 96.9), and placing it on psychotic-bipolar or metabolic liability was inadmissible. Under this model the insomnia pathway carried 56.3% of the sleep–internalizing covariance (95% CI 45.6–67.0%). A second sleep pathway improved fit (ΔAIC = −33.3), but with three sleep indicators its location was not identifiable, and across the equivalent models the insomnia share ranged from 0% to 127.0%. At the variant level, no internalizing lead departed from depression–anxiety proportionality after correction, and internalizing leads replicated in FinnGen, with partial sample overlap, whereas sleep leads did not (60.4%; P = 0.17). How concentrated the overlap is will require finer sleep phenotypes to establish. Among single-link models, insomnia is the best-supported location of the overlap.

## Introduction

Insomnia disorder affects 10–16% of adults and is an independent risk factor for other mental disorders [1]. People with insomnia who are not depressed have about twice the risk of later depression [2]. Whether insomnia acts as a cause, a consequence or a symptom of depression, or marks determinants the two share, is unresolved [3]. At the genetic level, genome-wide association studies (GWAS) of insomnia report genetic correlations with psychiatric traits [4,5], including depression [3,6], and anxiety disorders share much of their genetic liability with depression [7].

A genetic correlation shows that two traits share liability but not where within a domain that liability sits. For sleep and internalizing disorders, the spectrum of psychopathology that groups depressive and anxiety disorders [8], two accounts predict the same correlation. In a diffuse account, liability is shared between the domains as wholes, and each sleep phenotype relates to depression and anxiety in proportion to how well it reflects the sleep domain. In a concentrated account, part of the shared liability is specific to insomnia, the sleep complaint that is also a diagnostic criterion for major depressive disorder and generalized anxiety disorder [9]. A third account attributes the association to that shared diagnostic content rather than to shared liability. Which of them holds matters for research, because they point longitudinal, mechanistic and intervention studies to different sleep phenotypes. Separating them requires modelling several sleep and internalizing phenotypes at once.

Genomic structural equation modelling (GenomicSEM) fits such models to the genetic covariance among GWAS [10]. Because it allows a single indicator to load on two factors, the part of a between-domain relationship carried by that indicator can be estimated alongside the correlation between the factors. Applied to sleep health, GenomicSEM resolved six genetic factors, of which sleep duration, alertness and the absence of insomnia each related independently to an internalizing factor once their covariance was controlled [11,12]. Short and long sleep load on separate genetic factors [13], and insomnia, depression, anxiety and fibromyalgia share a common genetic factor [14]. Genetic studies of cardiometabolic and post-traumatic comorbidity have examined insomnia as a risk factor or mediator [15,16] and identified variants it shares with cardiometabolic disease [17]. None of these studies set diffuse, concentrated and shared-content accounts of the sleep–internalizing overlap against one another under a common rule.

We hypothesized that the sleep–internalizing overlap is concentrated in insomnia. If so, a link specific to insomnia should fit the genetic covariance better than links that span the sleep domain or represent shared diagnostic content. To test this, we fitted a four-factor model of sleep, internalizing, psychotic-bipolar and metabolic liability to ten GWAS. Starting from a model that links the two domains only through their factor correlation, we added in turn each cross-loading or shared-content residual covariance that could link them otherwise and compared the resulting accounts under one decision rule, with psychotic-bipolar and metabolic liability as comparators. We then asked how much of the cross-domain covariance runs through insomnia, and whether that quantity is identified. We also asked whether depression and anxiety hold together at the level of individual variants, as a spectrum model of their genetic risk implies [18], and whether the internalizing factor’s lead variants replicate in an independent biobank.

## Materials and methods

### Input GWAS and genetic covariance

Summary statistics for ten traits were obtained from public repositories that do not require a material transfer agreement (Table 1; Supplementary Table S1; workflow in Fig. 1a). The sleep traits were insomnia, sleep duration and chronotype from UK Biobank, through the MRC-IEU OpenGWAS resource [19]. The psychiatric traits were major depressive disorder (MDD) excluding 23andMe [20], anxiety disorders [7], bipolar disorder [21] and schizophrenia [22]. Anxiety disorders came from the public release of the PGC-ANX meta-analysis, which excludes the Utah cohort (122 083 cases and 729 602 controls), and schizophrenia from the public release of the primary multi-ancestry meta-analysis. The cardiometabolic traits were type 2 diabetes [23], body mass index (BMI) [24,25] and coronary artery disease [26]. Nine are of European ancestry, and all were analysed with a European linkage disequilibrium (LD) reference. Post-traumatic stress disorder (PTSD) entered only a post hoc sensitivity analysis, as a third internalizing indicator [27] (Supplementary Information).

**Figure 1.**
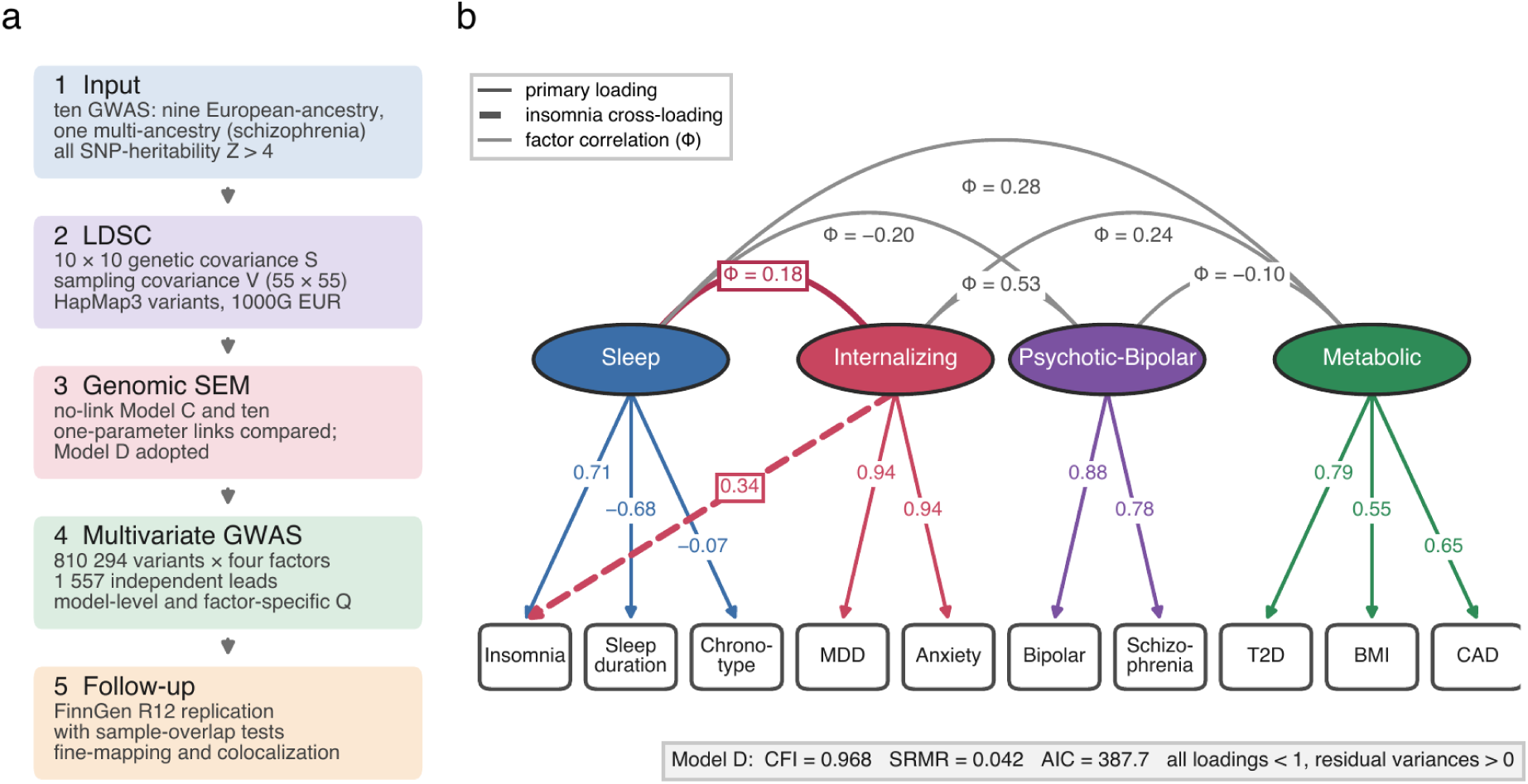
Study design and the adopted model. (**a**) Workflow. (**b**) Standardized solution of Model D. Ellipses are latent factors and rectangles the indicator GWAS. Solid arrows are primary loadings, the dashed arrow is the insomnia cross-loading, and curved lines are factor correlations (Φ), distinguished from LDSC genetic correlations between GWAS (r_g_). Fit and admissibility are given in the box. CAD, coronary artery disease; MDD, major depressive disorder; T2D, type 2 diabetes.

**Table 1.**
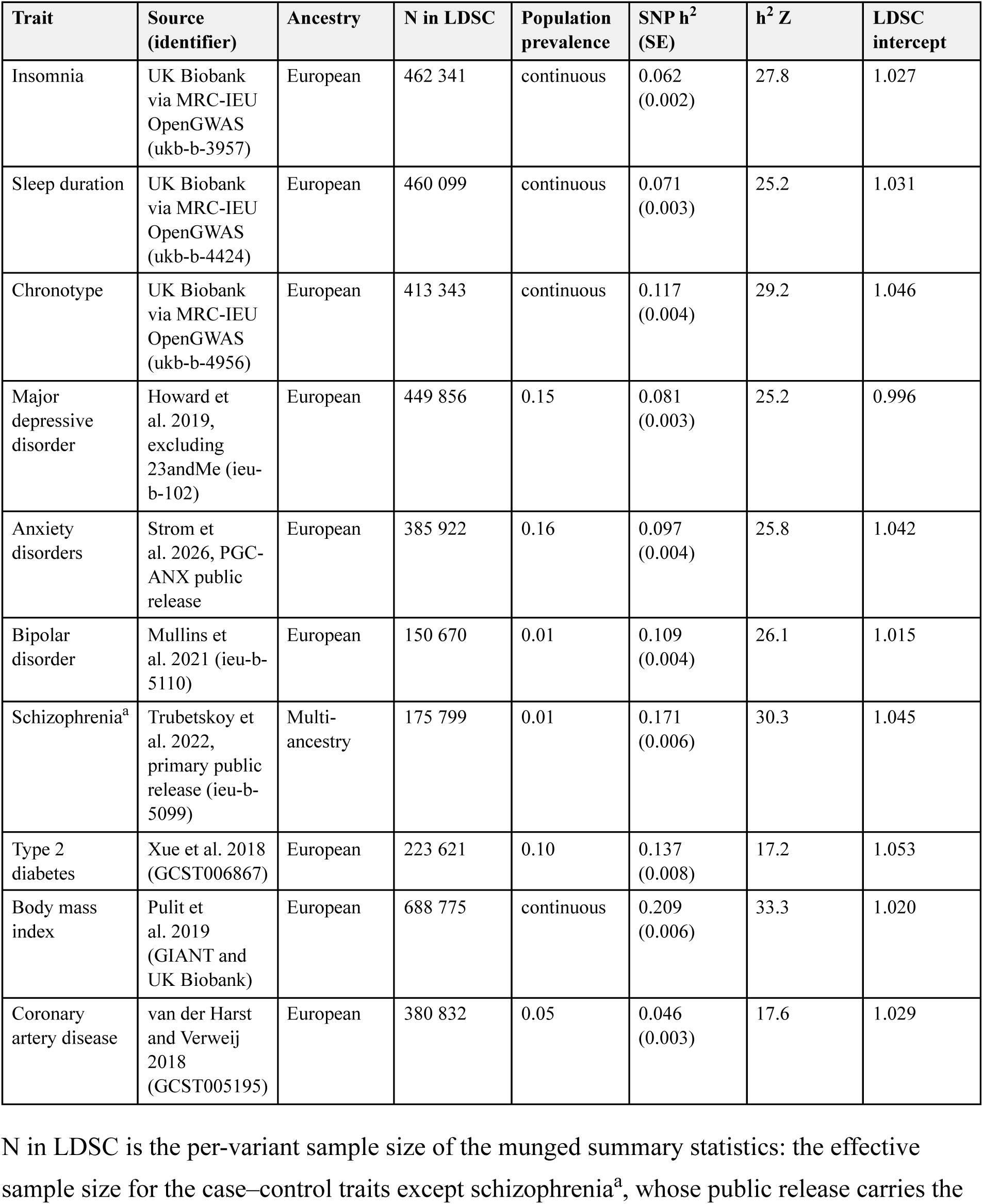
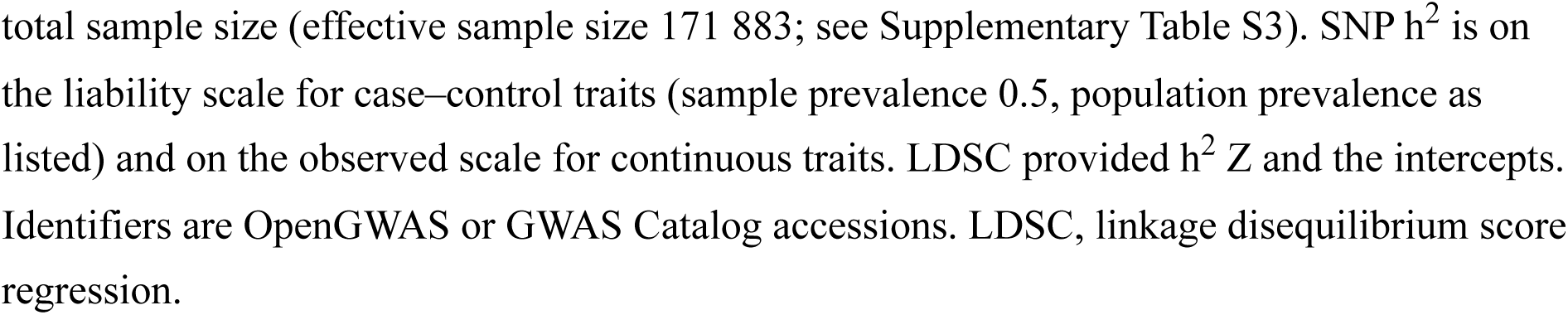
Input genome-wide association studies.

Summary statistics were munged in GenomicSEM against the HapMap3 variant list with a minor allele frequency threshold of 0.01 (Supplementary Information). SNP heritability (h^2^) and genetic covariance were estimated by LD score regression (LDSC) [28,29] in GenomicSEM, with European LD scores from 1000 Genomes Phase 3 [30] and HapMap3 variants [31], giving a 10 × 10 genetic covariance matrix and the sampling covariance of its elements (Supplementary Table S2). Each trait was required to reach an SNP-heritability Z statistic above 4. Case–control traits were converted to the liability scale with population prevalences of 0.15 (MDD), 0.16 (anxiety disorders), 0.01 (bipolar disorder and schizophrenia), 0.10 (type 2 diabetes) and 0.05 (coronary artery disease) and a sample prevalence of 0.5, since effective sample sizes were used. Because the schizophrenia file carries the total sample size and estimates of the prevalence of anxiety disorders vary, we refitted the model with the effective sample size for schizophrenia and with a prevalence of 0.20 for anxiety disorders (Supplementary Table S3). Throughout, r_g_ denotes an LDSC genetic correlation between two GWAS and Φ a correlation between latent factors.

### Factor models and the accounts compared

We fitted all models by diagonally weighted least squares in GenomicSEM [10], with factor variances fixed at 1. We treated a solution as admissible only if every standardized loading was at most 1 in absolute value and every residual variance was positive, and compared fit only among admissible solutions. Fit was summarized by the comparative fit index (CFI) and the standardized root mean square residual (SRMR), with CFI ≥ 0.95 and SRMR ≤ 0.08 as conventional adequacy [32], and by the Akaike information criterion (AIC). The root mean square error of approximation, which requires a single sample size, was not used (Supplementary Information).

The baseline structure, Model C, had four correlated factors: Sleep (insomnia, sleep duration, chronotype), Internalizing (MDD, anxiety disorders), Psychotic-Bipolar (bipolar disorder, schizophrenia) and Metabolic (type 2 diabetes, BMI, coronary artery disease), and we also fitted one- and two-factor models (Models A and B; Supplementary Table S4). In Model C the sleep and internalizing domains are linked only by their factor correlation, which is the diffuse account in its strictest form. We then added to Model C, in turn, each cross-loading between the two domains and each residual covariance representing shared diagnostic content, grouped by the account it represents (Table 2). Domain accounts keep the factor correlation as the carrier of the overlap and let one phenotype other than insomnia relate to the other domain beyond what that correlation implies: sleep duration or chronotype loading on Internalizing, or MDD or anxiety loading on Sleep. The insomnia account lets insomnia load on Internalizing (Model D). Shared-content accounts add a residual covariance between insomnia and MDD, between insomnia and anxiety, or both constrained to be equal, the last having as many parameters as Model D, and we also fitted a model with both covariances free. As comparators, the insomnia cross-loading was placed on Psychotic-Bipolar or Metabolic instead of Internalizing.

**Table 2.** One-parameter links added to the no-link model, comparators and two-link models.

| Account | Link(s) added to Model C | Admissible | $\chi^2$ (df) | $\Delta AIC^a$ | Max $\vert \lambda\vert $ | $\Phi$ | Insomnia share (%) |
| --- | --- | --- | --- | --- | --- | --- | --- |
| No link | none (factor correlation only) | no | 419.5 (29) | 83.9 | 1.197 | 0.368 | – |
| Insomnia account | insomnia → Internalizing (Model D) | yes | 333.7 (28) | 0 | 0.944 | 0.182 | 56.3 |
| Domain accounts | sleep duration → Internalizing | yes | 347.4 (28) | 13.8 | 0.943 | 0.552 | 0.0 |
|  | chronotype → Internalizing | no | 373.7 (28) | 40.0 | 1.151 | 0.384 | – |
|  | MDD → Sleep | no | 400.7 (28) | 67.0 | 1.186 | 0.326 | – |
|  | anxiety → Sleep | no | 400.7 (28) | 67.0 | 1.186 | 0.420 | – |
| Shared-content accounts | insomnia–MDD residual covariance | no | 411.0 (28) | 77.4 | 1.129 | 0.341 | – |
|  | insomnia–anxiety residual covariance | no | 411.0 (28) | 77.3 | 1.229 | 0.379 | – |
|  | both residual covariances, equal | yes | 430.6 (28) | 96.9 | 0.963 | 0.223 | 0.0 |
|  | both residual covariances, free<sup>b</sup> | yes | 427.0 (27) | 95.3 | 0.974 | 0.224 | 0.0 |
| Comparators | insomnia → Psychotic-Bipolar instead | no | 316.9 (28) | –16.8 | 1.073 | 0.250 | – |
|  | insomnia → Metabolic instead | no | 389.6 (28) | 55.9 | 1.994 | 0.331 | – |
| Two-link models | Model D + chronotype → Internalizing<sup>c</sup> | yes | 298.4 (27) | –33.3 | 0.939 | 0.202 | 49.6 |

| Account | Link(s) added to Model C | Admissible | $\chi^2$ (df) | $\Delta\text{AIC}^a$ | Max $\vert \lambda\vert $ | $\Phi$ | Insomnia share (%) |
| --- | --- | --- | --- | --- | --- | --- | --- |
|  | Model D + sleep duration → Internalizing<sup>c</sup> | yes | 298.4 (27) | −33.3 | 0.939 | −0.471 | 127.0 |
|  | sleep duration and chronotype → Internalizing<sup>c</sup> | yes | 298.4 (27) | −33.3 | 0.939 | 0.562 | 0.0 |
|  | Model D + MDD → Sleep | yes | 322.2 (27) | −9.5 | 0.965 | 0.135 | 55.2 |
|  | Model D + anxiety → Sleep | yes | 322.2 (27) | −9.5 | 0.984 | 0.238 | 56.3 |
|  | Model D + MDD → Metabolic | yes | 322.7 (27) | −9.0 | 0.958 | 0.175 | 55.9 |
|  | Model D + insomnia → Metabolic | yes | 323.6 (27) | −8.1 | 0.949 | 0.172 | 58.8 |
|  | Model D + insomnia → Psychotic-Bipolar | yes | 317.5 (27) | −14.2 | 0.995 | 0.236 | 36.6 |

Before fitting the rival models, we fixed the decision rule. Only admissible solutions were compared, and a ΔAIC below 2 was read as indistinguishable, 2 to 10 as preferred and above 10 as clearly preferred [33]. Because the AIC difference between two models fitted to one estimated matrix has sampling error, the competing models (listed in Supplementary Table S5) were refitted to 500 draws of the genetic covariance matrix from its LDSC sampling distribution, which gave the proportion of draws in which each account fitted better and an interval for each ΔAIC. Draws in which the Sleep factor emerged with all signs reversed were re-aligned (Supplementary Information). Finally, Model D was extended by each of seven second links, and a model loading both sleep duration and chronotype on Internalizing without an insomnia cross-loading was fitted, so that equivalent models, which fit identically while allocating parameters differently, could be identified [34]. Single-link models added to Model C cannot be transformed into one another without changing fit, which is why they can be compared by fit. We also refitted Model D with the residual variance of MDD, of anxiety disorders or of both fixed at 0.005, and ten times with one indicator omitted (Supplementary Tables S6 and S7). P values for added cross-loadings and for replication are not corrected for multiple comparisons.

### Covariance decomposition

Each model-implied genetic covariance between a sleep and an internalizing indicator is a sum of paths: through the factor correlation (λ_Sleep_ × Φ × λ_Internalizing_) and, in Model D, through the insomnia cross-loading (λ_insomnia,Internalizing_ × λ_Internalizing_). Because model-implied covariances can differ in magnitude and sign across the six pairs (three sleep by two internalizing indicators), the share carried by insomnia was summarized in three ways. The primary summary was the magnitude-weighted share, in which each pair contributes its insomnia share in proportion to the size of its covariance, so that it does not depend on cancellation between pairs of opposite sign. The other two were the net signed share, the summed insomnia paths divided by the summed covariances, and the unweighted average of the six per-pair shares (formulas in Supplementary Information). Intervals for the weighted and net shares came from the 500 Monte Carlo draws.

### Multivariate GWAS and variant-level heterogeneity

Under Model D, userGWAS was run on 810 294 HapMap3 variants, and variants that did not return estimates for all four factors were dropped. The extended major histocompatibility complex region and the sex chromosomes were not part of the analysed variant set. Independent leads for each factor were defined among variants with P < 5 × 10^−8^ by clumping in PLINK [35] at r^2^ < 0.1 within 500 kb, with a 1000 Genomes European reference. We assessed heterogeneity at the leads in three ways. The model-level Q_SNP_ compares the common-pathway model with one in which the variant acts on all ten indicators (6 df) [10]. Because it is model-level, a lead of one factor can be flagged for effects on another factor’s indicators. A factor-specific Q compares the common-pathway model with one in which the variant acts directly on the indicators of the factor in question while still acting on the other three factors [36], and for Internalizing these indicators include insomnia. Two 1-df tests asked whether a variant’s effect on MDD departs from the proportion implied by its effect on Internalizing, and likewise for bipolar disorder within Psychotic-Bipolar. Rates are reported at nominal significance, at a lead-set Bonferroni threshold (0.05 divided by the number of leads of the factor) and at a genome-wide Bonferroni threshold (0.05 / 810 294). To assess locus novelty, we lifted lead positions from GRCh37 to GRCh38, the build used by the GWAS Catalog [37], and computed the distance from each lead to the nearest catalogued association (May 2026 release) without trait filtering. A lead was classed as novel when no catalogued association lay within 100 kb.

### Replication in FinnGen and sample overlap

We tested lead variants in FinnGen release 12 [38] against an endpoint corresponding to one indicator of each factor: insomnia for Sleep, depression and anxiety for Internalizing, schizophrenia for Psychotic-Bipolar and type 2 diabetes for Metabolic. Alleles were harmonized to the factor effect allele, and each factor was oriented so that its highest-loading indicator loaded positively. We report direction concordance with a binomial test and the correlation of effect sizes. To read a factor’s concordance against that of its anchoring indicator, each indicator’s own genome-wide significant variants were clumped in the same way and tested against the same endpoint. Sample overlap between every input GWAS and every endpoint was estimated by the LDSC cross-trait intercept (Supplementary Information).

### Fine-mapping and colocalization

The two strongest leads of each factor were screened for mismatch with the out-of-sample LD reference (estimate_s_rss; bound 0.05) and fine-mapped with SuSiE [39] when they passed, with credible sets defined at 95% cumulative posterior probability. Colocalization with brain gene expression used full GTEx v8 summary statistics [40], uniformly reprocessed by the eQTL Catalogue [41], for cerebellar hemisphere and cerebellum, the two top-ranked tissues for Psychotic-Bipolar in tissue enrichment, and frontal cortex (BA9), the top-ranked tissue for Sleep and Internalizing (Supplementary Table S8). We used coloc.abf [42] under default priors and with p_12_ = 5 × 10^−6^ [43], and SuSiE-based colocalization with the eQTL Catalogue’s in-sample fine-mapping, which allows several causal signals per locus [44]. Because a factor GWAS has no sample size, analyses that require one used 500 199, the total sample size of the MDD GWAS, and fine-mapping was repeated across a 5.4-fold range, from 150 670, the smallest input effective sample size, to 810 294 (Supplementary Table S9). Single-variant colocalization used the factor effect sizes and standard errors with unit trait variance and does not depend on that value, and SuSiE-based colocalization used the fine-mapping at 500 199. Gene-based and tissue analyses with MAGMA [45,46] are reported in the Supplementary Information.

### Software and language model use

Analyses used R 4.4.3 with GenomicSEM 0.0.5, susieR 0.14.2 and coloc 5.2.3, and Python 3.11. A large language model (Claude, Anthropic) assisted with code development, numerical verification and language editing. The authors reviewed and verified all code, results and text and take responsibility for the content.

## Results

### Bivariate genetic correlations between the sleep and internalizing indicators

All ten traits had SNP-heritability Z between 17.2 and 33.3 and LDSC intercepts between 0.996 and 1.053 (Table 1). Insomnia was genetically correlated with MDD (r_g_ = 0.46, SE 0.03) and anxiety disorders (r_g_ = 0.40, SE 0.03). The corresponding correlations were −0.15 and −0.11 for sleep duration and 0.06 and 0.03 for chronotype (Fig. 2a; Supplementary Table S2). Within the sleep domain, insomnia and sleep duration correlated at −0.52, and chronotype correlated with insomnia at −0.03 and with sleep duration at 0.05. Within the internalizing domain, MDD and anxiety disorders correlated at 0.89. Neither scaling choice for schizophrenia or anxiety disorders changed the standardized model (Supplementary Table S3).

**Figure 2.**
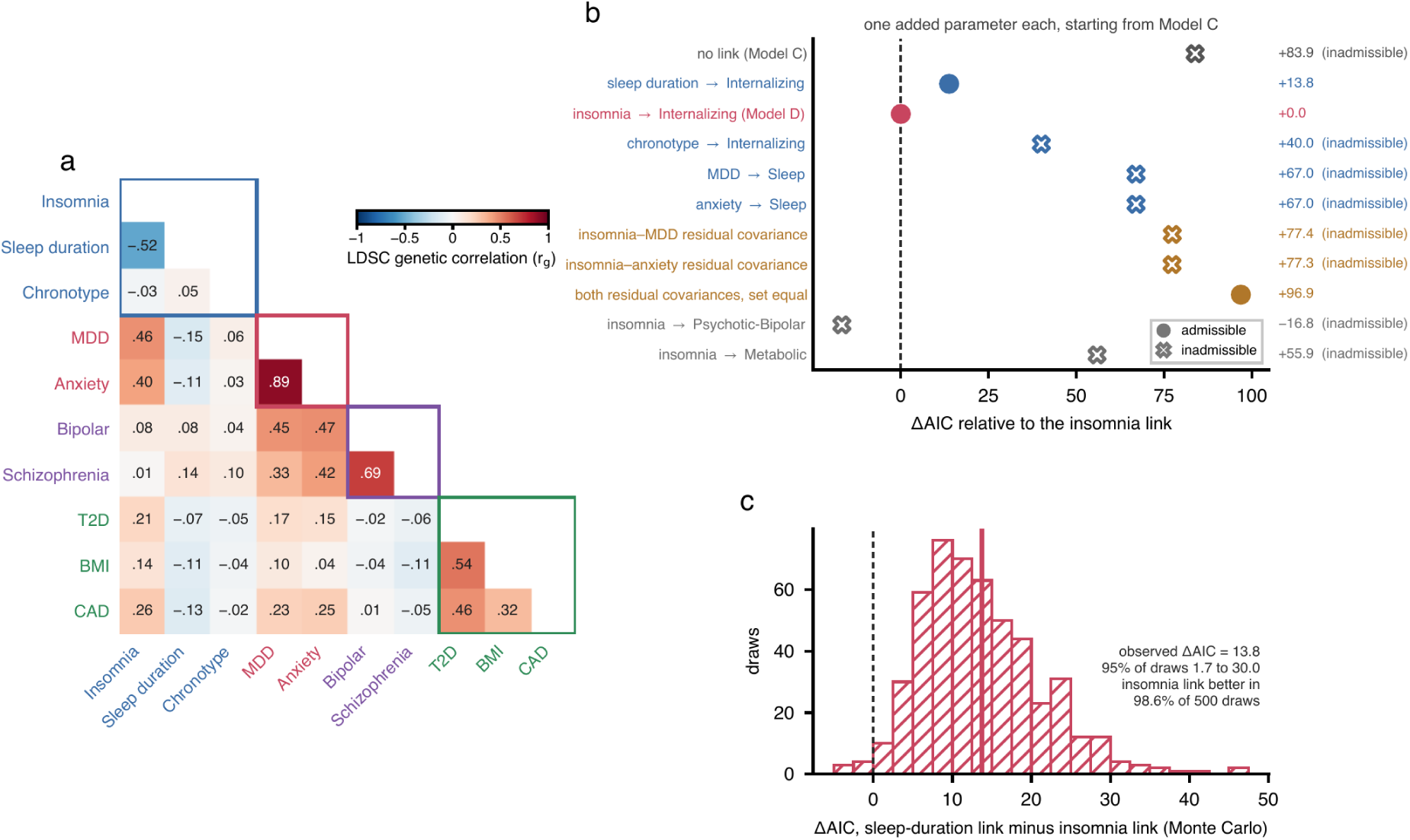
The no-link model and the single links that could repair it. (**a**) LDSC genetic correlations among the ten input GWAS. Labels are coloured by primary factor, and outlines mark each factor’s indicators. (**b**) ΔAIC relative to Model D for Model C and for each one-parameter link added to it, coloured by account (blue, domain; red, insomnia; orange, shared content; grey, comparators). Crosses mark inadmissible solutions, and values are listed on the right. (**c**) ΔAIC between the sleep-duration and insomnia links across 500 Monte Carlo draws of the genetic covariance matrix. The solid line marks the observed value and the dashed line zero.

### The no-link model is inadmissible, and the insomnia link repairs it best

One- and two-factor models had CFI = 0.476 and 0.639, below the adequacy threshold. Model C met conventional adequacy (CFI = 0.960, SRMR = 0.048) but was inadmissible: the standardized loading of insomnia on Sleep was 1.197 and its residual variance −0.432 (Table 2).

Of the ten one-parameter repairs, three were admissible: the insomnia cross-loading, a sleep-duration cross-loading and the pair of equal residual covariances (Fig. 2b; Table 2). In the insomnia account (Model D), insomnia loaded on Sleep at 0.706 (95% CI 0.636–0.777) and on Internalizing at 0.340 (0.290–0.391), and the factor correlation was Φ = 0.182 (0.122–0.242). Fit was CFI = 0.968, SRMR = 0.042 and AIC = 387.7 (Fig. 1b; Supplementary Table S10). In the same model, Sleep correlated with Metabolic at Φ = 0.282 without a cross-loading, and Internalizing with Psychotic-Bipolar at Φ = 0.532 (Fig. 1b; Supplementary Table S10). In the only admissible domain account, sleep duration loaded on Internalizing (0.302) as well as on Sleep (−0.783), the factor correlation rose to Φ = 0.552, and insomnia loaded on Sleep at 0.850. The insomnia account fitted better (ΔAIC = 13.8), clearly preferred under the decision rule. It fitted better in 98.6% of the 500 Monte Carlo draws, with ΔAIC between 1.7 and 30.0 in 95% of them and above 10 in 63.6% (Fig. 2c).

Loading chronotype on Internalizing, or MDD or anxiety on Sleep, left the solution inadmissible, as did a single residual covariance between insomnia and either internalizing indicator. With both residual covariances, constrained to be equal so as to match Model D in complexity, the solution was admissible and fitted worse (ΔAIC = 96.9; worse in all 500 draws). Placing the insomnia cross-loading on Psychotic-Bipolar instead of Internalizing lowered AIC (ΔAIC = −16.8) but left the insomnia loading on Sleep above 1 (1.073), and the solution was admissible in only 4.2% of draws. On Metabolic it was inadmissible in all 464 draws that converged.

### About half of the cross-domain covariance runs through insomnia

Under Model D, the two insomnia pairs carried 72.5% of their covariance through the cross-loading, and the four pairs involving sleep duration and chronotype (covariances −0.116 to −0.011) carried none by construction (Fig. 3a; Supplementary Table S11). Weighted by magnitude, the insomnia pathway carried 56.3% of the sleep–internalizing covariance (95% CI 45.6–67.0%), more than half in 86.8% of draws. The net signed share was 101.9% (93.2– 110.7%): the factor-correlation paths of the negatively loading sleep indicators cancel those of insomnia, leaving almost all of the net covariance to the cross-loading. The unweighted average of the six per-pair shares was 24.2%, because four of the six pairs carry no insomnia path by construction. With PTSD added as a third internalizing indicator, the insomnia account was again preferred over the domain account (ΔAIC = 12.5), and the weighted share was 51.1% (Supplementary Table S12).

**Figure 3.**
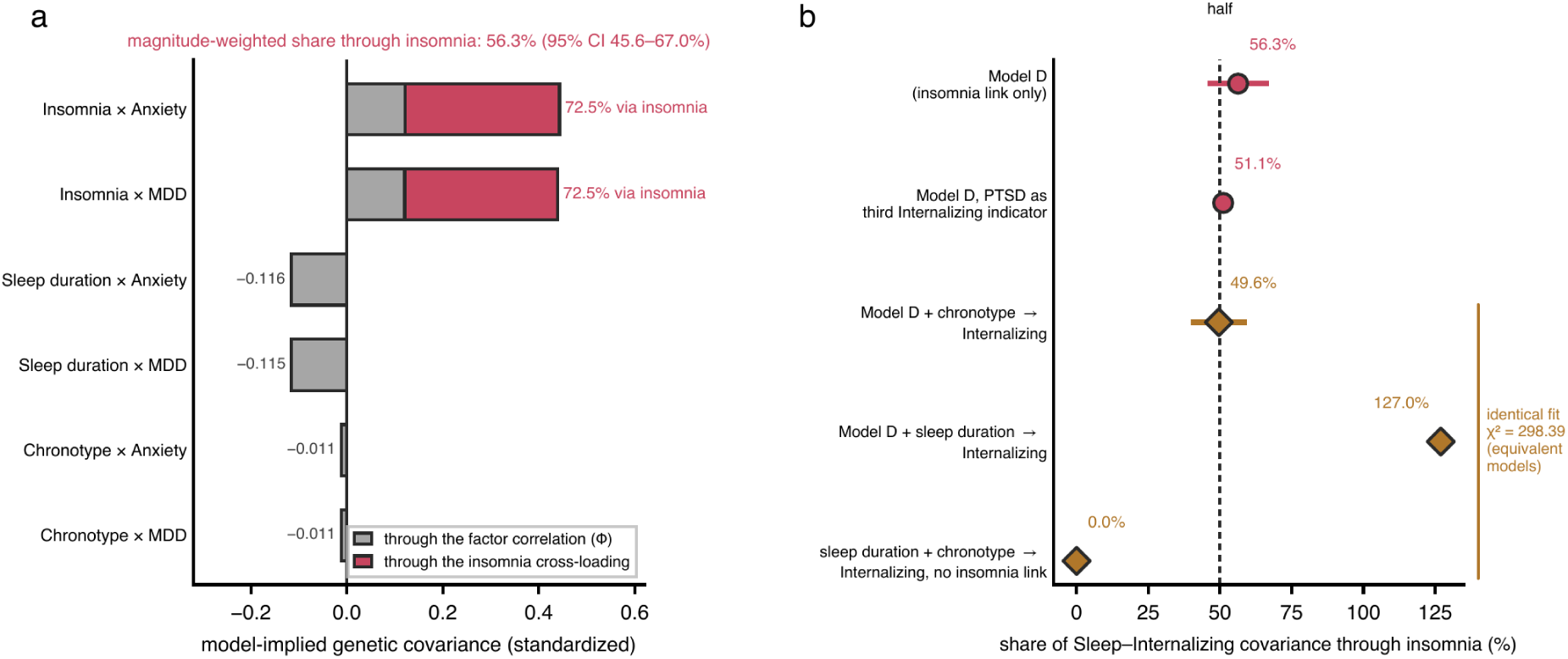
How much of the cross-domain covariance runs through insomnia. (**a**) Model-implied genetic covariance of the six Sleep × Internalizing indicator pairs under Model D, divided into the part carried by the factor correlation (grey) and by the insomnia cross-loading (red). The magnitude-weighted share and its Monte Carlo 95% interval are given above. (**b**) Share carried by insomnia under Model D, under Model D with post-traumatic stress disorder as a third internalizing indicator, and under the three equivalent two-link models (diamonds), which fit identically. Horizontal bars are Monte Carlo 95% intervals where computed.

### With a second sleep pathway, the location of the overlap is not identified

Each of the seven second links added to Model D improved fit (Supplementary Table S13). The largest improvement came from a chronotype cross-loading on Internalizing (0.081; ΔAIC = −33.3, between −66.1 and −10.6 in 95% of draws), under which the weighted insomnia share was 49.6% (39.8–59.5%). This model is one of three that fit identically (χ^2^ = 298.39 on 27 df; Table 2). The same improvement follows from adding a sleep-duration rather than a chronotype cross-loading to Model D, which reverses the factor correlation (Φ = −0.471) and raises the insomnia share to 127.0%, or from dropping the insomnia cross-loading and loading both sleep duration and chronotype on Internalizing (Φ = 0.562), which reduces it to 0% (Fig. 3b). The three are equivalent because the Sleep factor can be redefined as a weighted combination of itself and the Internalizing factor without changing fit. Each redefinition moves the cross-loadings among the sleep indicators, and with two cross-loadings any of the three indicators can be the one left without [34]. The same equivalence held with PTSD included (Supplementary Table S12).

Adding MDD or anxiety on Sleep (ΔAIC = −9.5 for either) or MDD on Metabolic (−9.0) left the insomnia cross-loading at 0.335–0.342, and an added insomnia cross-loading on Metabolic was 0.096 (P = 0.003; ΔAIC = −8.1) and left the Internalizing cross-loading at 0.330. An added cross-loading on Psychotic-Bipolar lowered AIC by 14.2. In that model the loading on Internalizing fell to 0.075 (P = 0.63), that on Psychotic-Bipolar was 0.275 (P = 0.096) and that on Sleep 0.995, and the standard errors of insomnia’s loadings rose five-to sixfold. Eight of the ten leave-one-indicator-out refits were admissible, and in these the cross-loading stayed between 0.33 and 0.37 whenever sleep duration was retained, and rose to 0.60 without it (Supplementary Table S7). Fixing the residual variances of the internalizing indicators left it at 0.33–0.34 (Supplementary Table S6).

### The internalizing factor is coherent at the level of variants

Of the 810 294 variants, 810 109 returned estimates for all four factors, and 1 557 independent leads reached genome-wide significance: 53 for Sleep, 147 for Internalizing, 255 for Psychotic-Bipolar and 1 102 for Metabolic (Supplementary Table S14; Supplementary Fig. S1). λ_GC_ was 1.56–2.84, LDSC intercepts 1.014–1.187 and attenuation ratios 0.020–0.064 (Supplementary Table S15). Every lead lay within 100 kb of an association already in the GWAS Catalog (Supplementary Table S16).

On the model-level Q_SNP_, 45.2–56.6% of each factor’s leads were heterogeneous at nominal significance, 9.4–26.4% at the lead-set Bonferroni threshold and 3.5–9.4% at the genome-wide threshold (Fig. 4a; Supplementary Table S17). On the factor-specific Q, 2.7% of Internalizing, 3.5% of Metabolic, 4.3% of Psychotic-Bipolar and 18.9% of Sleep leads were heterogeneous at the lead-set threshold. In the 1-df test, no Internalizing lead departed from MDD–anxiety proportionality after correction (0 of 147; 11.6% at nominal significance), whereas 4.3% of Psychotic-Bipolar leads departed from bipolar disorder–schizophrenia proportionality.

**Figure 4.**
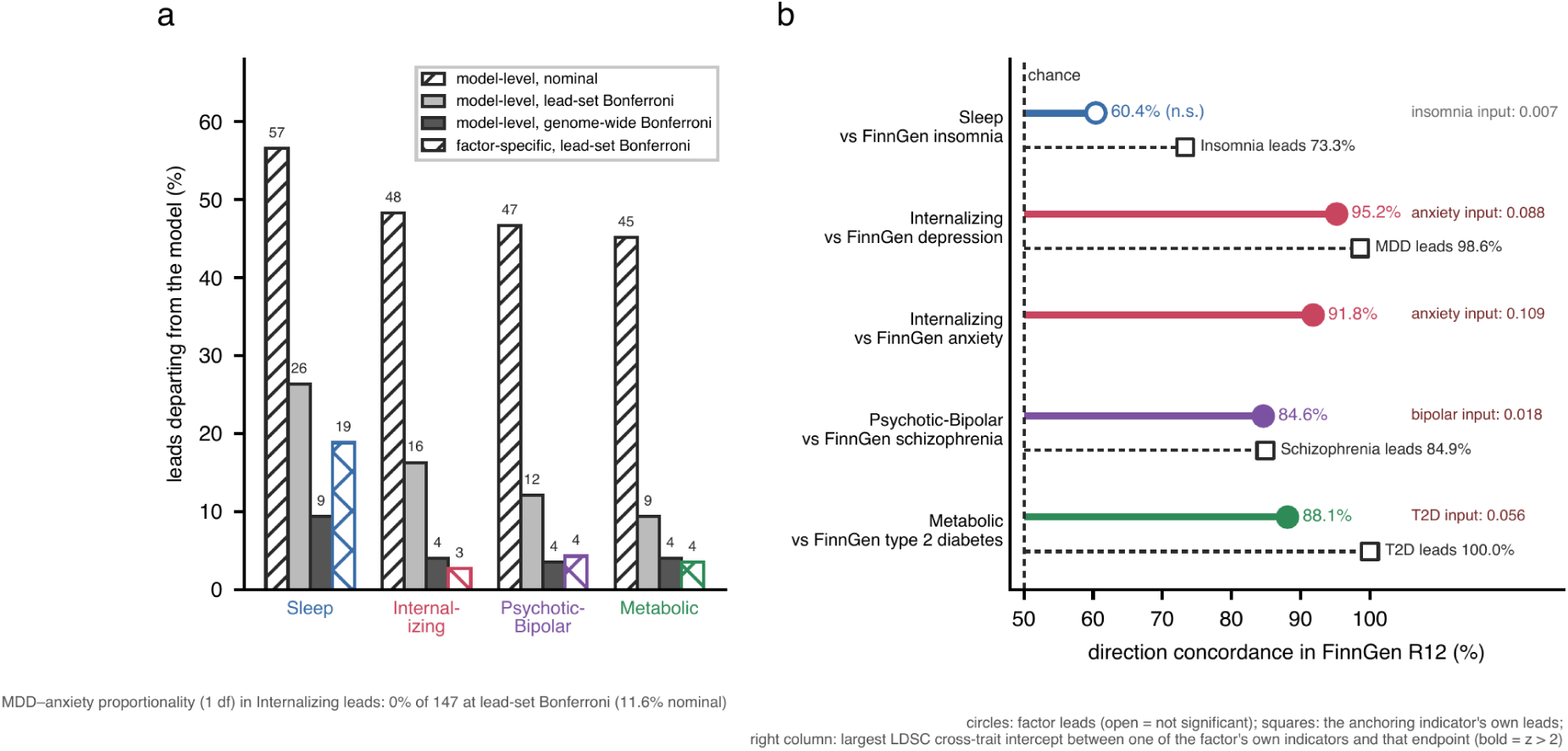
Variant-level heterogeneity and replication. (a) Percentage of each factor’s lead variants departing from the model on the model-level Q_SNP_ at three thresholds and on the factor-specific Q at the lead-set Bonferroni threshold. The 1-df test of MDD–anxiety proportionality is summarized below the panel. (**b**) Direction concordance in FinnGen R12 of factor leads (circles; open when not significant) and of each anchoring indicator’s own leads (squares). The right column gives the largest LDSC cross-trait intercept between one of the factor’s own indicators and the endpoint, in bold when z > 2.

### Direction concordance of lead variants in FinnGen

In FinnGen R12, direction concordance of factor leads was 88.1% for Metabolic against type 2 diabetes (1 096 of 1 102 leads matched; P = 2.2 × 10^−158^; r = 0.72), 84.6% for Psychotic-Bipolar against schizophrenia (254 of 255; P = 1.2 × 10^−30^; r = 0.71), 95.2% and 91.8% for Internalizing against depression and anxiety (147 of 147; P = 3.0 × 10^−33^ and 1.6 × 10^−27^; r = 0.86 and 0.83) and 60.4% for Sleep against insomnia (53 of 53; P = 0.17; r = 0.28) (Fig. 4b; Supplementary Table S18). The anchoring indicators’ own leads were concordant at 98.6% for MDD (72 of 74; P = 3.1 × 10^−20^; r = 0.86), 84.9% for schizophrenia (351 of 355; P = 1.6 × 10^−42^; r = 0.76), 100.0% for type 2 diabetes (173 of 173; P = 1.7 × 10^−52^; r = 0.96), 73.3% for insomnia (60 of 60; P = 3.9 × 10^−4^; r = 0.72) and 50.0% for sleep duration (102 of 105; P = 1.0; r = 0.10) (Supplementary Table S19). Cross-trait intercepts were 0.109 and 0.088 between the anxiety input and the FinnGen anxiety and depression endpoints, 0.056 between the type 2 diabetes input and its endpoint and 0.018 between the bipolar input and the schizophrenia endpoint, and the other inputs had intercepts between −0.008 and 0.010 with the endpoint of their own factor (Supplementary Table S20).

### Locus follow-up

Four of the eight strongest leads passed the LD-mismatch screen and were fine-mapped, and the fine-mapping results were unchanged across the range of assumed sample sizes (Supplementary Tables S9 and S21; Supplementary Fig. S2). With full eQTL summary statistics, the Internalizing locus on chromosome 1 (rs3101336) colocalized with expression of the long non-coding RNA LINC02796 (ENSG00000286863 in Ensembl release 111) in frontal cortex and cerebellar hemisphere (PP.H4 ≥ 0.99), but not with *NEGR1* (PP.H4 ≤ 0.30). At *CACNA1C* (rs4298967, Psychotic-Bipolar), single-variant colocalization favoured distinct causal variants, whereas SuSiE-based colocalization matched one of the two GWAS signals to a *CACNA1C* eQTL signal in cerebellar hemisphere (PP.H4 = 0.99) and cerebellum (0.71) (Supplementary Table S22).

## Discussion

Of the single links that could connect sleep and internalizing liability, an insomnia-specific cross-loading fitted best, and under it 56.3% (95% CI 45.6–67.0%) of the cross-domain covariance ran through insomnia. A factor correlation alone could not carry insomnia’s covariance with the internalizing indicators without implying a sleep factor that insomnia measures more than perfectly, so the strictly diffuse account fails. The insomnia link was preferred over the only admissible domain-wide alternative consistently in direction across Monte Carlo draws, though less consistently in size, and over residual covariances representing shared diagnostic content in every draw. This refines the insomnia–depression genetic correlation, because in the preferred model insomnia relates to internalizing liability more strongly than its loading on the sleep factor predicts.

Mendelian randomization studies have suggested that liability to insomnia increases the risk of depression [4,5], and a genomic factor model placed insomnia, depression, anxiety and fibromyalgia on one common factor [14]. Our model adds structure to this evidence, because insomnia loaded on the sleep factor and also related to internalizing liability through a specific path, an arrangement that fitted better than domain-wide and shared-content alternatives, which had not previously been compared under one rule.

The two admissible cross-loading accounts have the same form, each adding one phenotype-specific cross-loading to Model C, and differ in what carries the overlap. Under the sleep-duration link the factor correlation rises from 0.18 to 0.55 and does most of the carrying. Under the insomnia link it stays small and a specific path carries about half, and the data favour this description.

The comparators bound the specificity of this path. Sleep’s association with Metabolic needed no cross-loading, and an insomnia cross-loading onto Metabolic added to Model D was less than a third of the Internalizing cross-loading, which stayed at 0.33. Psychotic-Bipolar, which correlates with Internalizing at Φ = 0.53, could not be separated from it. Once insomnia was allowed to load on both, the two cross-loadings shared the covariance and neither was distinguishable from zero. The data thus distinguish the internalizing target from the metabolic one but not from the psychotic-bipolar one.

The data also favour a second sleep pathway, but with three sleep indicators its location cannot be identified, and once it is admitted the share attributable to insomnia ranges from 0% to 127.0% across models that fit identically. The data therefore identify insomnia as the best-supported single location of the overlap, but not the fraction it carries. More generally, whenever cross-loadings can move between indicators of a factor, a decomposition of between-factor covariance rests on an identification choice, and the equivalent models should be reported with the adopted one [34].

These results converge with earlier genomic work on sleep. In the six-factor sleep health model, duration and alertness as well as the absence of insomnia related independently to internalizing liability [11,12]. Here, too, a second sleep pathway improved fit, and sleep duration provided the only admissible domain-wide link. Sleep duration has a genetic architecture largely distinct from other sleep measures, and short and long sleep load on separate factors [13]. That may be one reason a single duration indicator cannot locate the second pathway: pathways carried by short and by long sleep could offset each other in a linear measure of duration.

The insomnia pathway outperformed residual covariances representing shared diagnostic content, which addresses a long-standing concern. Insomnia is a criterion for both depression and generalized anxiety disorder [9], so its genetic overlap with them might reflect how these disorders are defined. The residual covariances were inadmissible singly and, jointly, fitted worse than the cross-loading in every Monte Carlo draw. Overlapping item content may still inflate the size of the insomnia pathway, but it does not account for its presence.

At the variant level the internalizing factor behaved as one dimension: no lead departed from MDD–anxiety proportionality after correction. Internalizing is an established transdiagnostic dimension in phenotypic and twin research [47], and genomic studies have asked whether such dimensions hold when genetic risk is examined directly [48,49]. Our result supports modelling depression and anxiety as indicators of shared liability, as dimensional taxonomies do [8,18], with the qualification that the dimension estimated here is the liability shared by these two disorders, not the broader internalizing spectrum. The sleep factor was the least coherent, at the variant level as in FinnGen, consistent with indicators that measure distinct phenotypes: insomnia leads replicated against a clinical insomnia endpoint, and sleep-duration leads did not. Inflation at the factor level was largely polygenic, and every lead lay in a region already associated in the GWAS Catalog, so the contribution of the factor GWAS is structural rather than one of locus discovery.

At the locus level, colocalization implicated *CACNA1C*, a cross-disorder locus [50], in cerebellar tissue for Psychotic-Bipolar, but only once the two association signals at the locus were modelled separately. A single-signal analysis favoured distinct causal variants. The cerebellum contributes to emotional processing [51] and to cortico-cerebellar models of schizophrenia [52], and earlier *CACNA1C* work emphasized the prefrontal cortex [53]. We nominate the locus–tissue pair for replication rather than reading mechanism into it, and we note that the colocalization at the chromosome 1 Internalizing locus was with the long non-coding RNA LINC02796, not with *NEGR1*.

The results suggest two hypotheses for clinical research. Studies that use sleep to predict, explain or prevent internalizing disorders should treat insomnia as the primary sleep phenotype, rather than composite sleep measures that combine insomnia with phenotypes whose share of the overlap is not identified. And because the internalizing factor is coherent across depression and anxiety, prevention trials of insomnia treatment should measure both. Interventions that improve sleep reduce depressive and anxiety symptoms by similar margins, with pooled effects of g = −0.63 and −0.51 across 65 randomized trials [54] and of −0.36 and −0.35 for internet-delivered cognitive behavioural therapy for insomnia [55]. Of the prevention and early-intervention trials of cognitive behavioural therapy for insomnia in a recent meta-analysis, seven examined depressive symptoms and one an anxiety disorder [56].

### Limitations

FinnGen replication is partly internal for Internalizing, because the anxiety input shares participants with FinnGen, and it tested lead variants against single endpoints, so the factor structure itself has not been reproduced in an independent sample. Nine inputs were of European ancestry, and the schizophrenia input was multi-ancestry but analysed with a European LD reference. For several traits, larger GWAS than those used here are now available [57–60], and newer releases could change the indicator loadings. The internalizing factor rests on two indicators, and adding PTSD as a third moved the insomnia share from 56% to 51%. The three sleep indicators are single self-report items from UK Biobank, where reporting error attenuates genome-wide estimates [61], and the insomnia indicator is a symptom item rather than a diagnosis. Genetic covariance does not establish causal direction. The rival accounts were specified after Model D and its two-link extensions had been fitted, and the decision rule was set before the rival models were fitted. Because the rival set comprises every cross-loading between the two domains and the residual covariances representing shared diagnostic content, its later specification could not favour one account.

Fine-mapping relied on an out-of-sample LD reference and on an assumed sample size for the factor GWAS, colocalization covered three brain tissues and its posteriors depend on the priors, and the extended major histocompatibility complex region and the sex chromosomes were not analysed.

## Conclusion

Among single-link models, insomnia is the best-supported location of the genetic overlap between sleep and internalizing liability: it is preferred over domain-wide and shared-content accounts and carries about half of the cross-domain covariance. How concentrated the overlap is cannot be determined with three sleep indicators. Modelling short and long sleep, daytime sleepiness and device-based measures of sleep as separate indicators could make the second pathway identifiable, and separating the shared and sleep-specific components of insomnia liability at the level of variants could show which loci carry the overlap.

## Supporting information

Supplementary Information (methods, results, Figures S1-S2)

## Data Availability

All input GWAS summary statistics are publicly available, and accession identifiers or download sources are listed in Table 1. FinnGen R12 endpoint statistics, GTEx v8 eQTL summary statistics through the eQTL Catalogue and the GWAS Catalog are available through their portals. Factor-level GWAS summary statistics will be deposited in the GWAS Catalog on publication. The analysis code is available from the corresponding author on request and will be deposited in a public repository with a persistent DOI on publication.

https://gwas.mrcieu.ac.uk

https://pgc.unc.edu

https://www.ebi.ac.uk/gwas

https://www.finngen.fi/en/access_results

https://www.ebi.ac.uk/eqtl

## Acknowledgements

We thank the participants and investigators of UK Biobank, the Psychiatric Genomics Consortium, DIAGRAM, GIANT and CARDIoGRAMplusC4D for making their summary statistics publicly available, and the MRC Integrative Epidemiology Unit at the University of Bristol for the OpenGWAS resource. We want to acknowledge the participants and investigators of the FinnGen study. The Genotype-Tissue Expression (GTEx) Project was supported by the Common Fund of the Office of the Director of the National Institutes of Health, and by NCI, NHGRI, NHLBI, NIDA, NIMH, and NINDS, and GTEx v8 expression data were obtained through the eQTL Catalogue. The numerical calculations reported in this paper were partially performed at TUBITAK ULAKBIM, High Performance and Grid Computing Center (TRUBA resources).

## Funding

This research received no specific grant from any funding agency in the public, commercial or not-for-profit sectors.

## Conflict of interest

The authors declare no competing interests.

## Author contributions

Mehmet Akif Şakiroğlu: conceptualization, methodology, software, formal analysis, investigation, visualization, writing – original draft. Yusuf Çiçek: investigation, software, validation, writing – review and editing. Murat Can Çelik: investigation, validation, writing – review and editing. Ali Tarık Altunç: investigation, validation, writing – review and editing. Doğukan Aslan: investigation, writing – review and editing. Halil Aziz Velioğlu: conceptualization, supervision, project administration, writing – review and editing. All authors read and approved the final manuscript.

## Ethics approval and consent to participate

This study analysed publicly available, de-identified genome-wide association summary statistics, and no new data were collected from human participants. Ethical approval and informed consent were obtained by the original studies, which are cited in the Methods and listed in Table 1.

## Data availability

All input GWAS summary statistics are publicly available, and accession identifiers or download sources are listed in Table 1. FinnGen R12 endpoint statistics, GTEx v8 eQTL summary statistics through the eQTL Catalogue and the GWAS Catalog are available through their portals. Factor-level GWAS summary statistics will be deposited in the GWAS Catalog on publication.

## Code availability

The analysis code is available from the corresponding author on request and will be deposited in a public repository with a persistent DOI on publication.

## Notes

### Competing Interest Statement

The authors have declared no competing interest.

### Author Declarations

MRC IEU OpenGWAS, the Psychiatric Genomics Consortium, the GWAS Catalog, FinnGen release 12 and the eQTL Catalogue (GTEx v8). Accession identifiers are listed in Table 1 of the manuscript.

## References

1. Spiegelhalder K, et al. Insomnia disorder. Nat Rev Dis Primers 2026; 12: 17.

2. Baglioni C et al. Insomnia as a predictor of depression: a meta-analytic evaluation of longitudinal epidemiological studies. J Affect Disord 2011; 135: 10–19.

3. Kozhemiako N et al. The interplay between sleep and mental health: a genetic perspective. Biol Psychiatry 2025; 98: 863–874.

4. Jansen PR et al. Genome-wide analysis of insomnia in 1,331,010 individuals identifies new risk loci and functional pathways. Nat Genet 2019; 51: 394–403.

5. Lane JM et al. Biological and clinical insights from genetics of insomnia symptoms. Nat Genet 2019; 51: 387–393.

6. Stein MB et al. Genome-wide analysis of insomnia disorder. Mol Psychiatry 2018; 23: 2238–2250.

7. Strom NI et al. Genome-wide association study of major anxiety disorders in 122,341 European-ancestry cases identifies 58 loci and highlights GABAergic signaling. Nat Genet 2026; 58: 275–288.

8. Kotov R et al. The Hierarchical Taxonomy of Psychopathology (HiTOP): a dimensional alternative to traditional nosologies. J Abnorm Psychol 2017; 126: 454–477.

9. American Psychiatric Association. Diagnostic and Statistical Manual of Mental Disorders, 5th edn (American Psychiatric Publishing, Arlington, VA, 2013).

10. Grotzinger AD et al. Genomic structural equation modelling provides insights into the multivariate genetic architecture of complex traits. Nat Hum Behav 2019; 3: 513–525.

11. Morrison CL et al. Sleep health at the genomic level: six distinct factors and their relationships with psychopathology. Biol Psychiatry Glob Open Sci 2023; 3: 530–540.

12. Morrison CL, Winiger EA, Wright KP, Friedman NP. Multivariate genome-wide association study of sleep health demonstrates unity and diversity. Sleep 2024; 47: zsad320.

13. Austin-Zimmerman I et al. Multi-ancestry genetic architecture of sleep duration and its relationship to other sleep and psychiatric phenotypes. Preprint at medRxiv 10.1101/2025.05.19.25327902 (2025).

14. Lin L et al. Shared latent genetic liability across fibromyalgia and psychiatric traits: novel insights from genomic structural equation modeling. PLoS Genet 2026; 22: e1012034.

15. Baltramonaityte V et al. A multivariate genome-wide association study of psycho-cardiometabolic multimorbidity. PLoS Genet 2023; 19: e1010508.

16. Lukas E et al. A genetic exploration of the relationship between posttraumatic stress disorder and cardiovascular diseases. Transl Psychiatry 2025; 15: 1.

17. Zhang Y et al. Shared genetic architecture between sleep traits and cardiometabolic diseases. EBioMedicine 2026; 126: 106220.

18. Waszczuk MA et al. Redefining phenotypes to advance psychiatric genetics: implications from hierarchical taxonomy of psychopathology. J Abnorm Psychol 2020; 129: 143–161.

19. Elsworth B et al. The MRC IEU OpenGWAS data infrastructure. Preprint at bioRxiv 10.1101/2020.08.10.244293 (2020).

20. Howard DM et al. Genome-wide meta-analysis of depression identifies 102 independent variants and highlights the importance of the prefrontal brain regions. Nat Neurosci 2019; 22: 343–352.

21. Mullins N et al. Genome-wide association study of more than 40,000 bipolar disorder cases provides new insights into the underlying biology. Nat Genet 2021; 53: 817–829.

22. Trubetskoy V et al. Mapping genomic loci implicates genes and synaptic biology in schizophrenia. Nature 2022; 604: 502–508.

23. Xue A et al. Genome-wide association analyses identify 143 risk variants and putative regulatory mechanisms for type 2 diabetes. Nat Commun 2018; 9: 2941.

24. Pulit SL et al. Meta-analysis of genome-wide association studies for body fat distribution in 694 649 individuals of European ancestry. Hum Mol Genet 2019; 28: 166–174.

25. Locke AE et al. Genetic studies of body mass index yield new insights for obesity biology. Nature 2015; 518: 197–206.

26. van der Harst P, Verweij N. Identification of 64 novel genetic loci provides an expanded view on the genetic architecture of coronary artery disease. Circ Res 2018; 122: 433– 443.

27. Nievergelt CM et al. Genome-wide association analyses identify 95 risk loci and provide insights into the neurobiology of post-traumatic stress disorder. Nat Genet 2024; 56: 792–808.

28. Bulik-Sullivan BK et al. LD score regression distinguishes confounding from polygenicity in genome-wide association studies. Nat Genet 2015; 47: 291–295.

29. Bulik-Sullivan B et al. An atlas of genetic correlations across human diseases and traits. Nat Genet 2015; 47: 1236–1241.

30. 1000 Genomes Project Consortium. A global reference for human genetic variation. Nature 2015; 526: 68–74.

31. International HapMap 3 Consortium. Integrating common and rare genetic variation in diverse human populations. Nature 2010; 467: 52–58.

32. Hu L, Bentler PM. Cutoff criteria for fit indexes in covariance structure analysis: conventional criteria versus new alternatives. Struct Equ Modeling 1999; 6: 1–55.

33. Burnham KP, Anderson DR. Multimodel inference: understanding AIC and BIC in model selection. Sociol Methods Res 2004; 33: 261–304.

34. MacCallum RC, Wegener DT, Uchino BN, Fabrigar LR. The problem of equivalent models in applications of covariance structure analysis. Psychol Bull 1993; 114: 185– 199.

35. Chang CC et al. Second-generation PLINK: rising to the challenge of larger and richer datasets. Gigascience 2015; 4: 7.

36. Grotzinger AD et al. Genetic architecture of 11 major psychiatric disorders at biobehavioral, functional genomic and molecular genetic levels of analysis. Nat Genet 2022; 54: 548–559.

37. Sollis E et al. The NHGRI-EBI GWAS Catalog: knowledgebase and deposition resource. Nucleic Acids Res 2023; 51: D977–D985.

38. Kurki MI et al. FinnGen provides genetic insights from a well-phenotyped isolated population. Nature 2023; 613: 508–518.

39. Zou Y, Carbonetto P, Wang G, Stephens M. Fine-mapping from summary data with the “Sum of Single Effects” model. PLoS Genet 2022; 18: e1010299.

40. GTEx Consortium. The GTEx Consortium atlas of genetic regulatory effects across human tissues. Science 2020; 369: 1318–1330.

41. Kerimov N et al. A compendium of uniformly processed human gene expression and splicing quantitative trait loci. Nat Genet 2021; 53: 1290–1299.

42. Giambartolomei C et al. Bayesian test for colocalisation between pairs of genetic association studies using summary statistics. PLoS Genet 2014; 10: e1004383.

43. Wallace C. Eliciting priors and relaxing the single causal variant assumption in colocalisation analyses. PLoS Genet 2020; 16: e1008720.

44. Wallace C. A more accurate method for colocalisation analysis allowing for multiple causal variants. PLoS Genet 2021; 17: e1009440.

45. de Leeuw CA, Mooij JM, Heskes T, Posthuma D. MAGMA: generalized gene-set analysis of GWAS data. PLoS Comput Biol 2015; 11: e1004219.

46. Frankish A et al. GENCODE reference annotation for the human and mouse genomes. Nucleic Acids Res 2019; 47: D766–D773.

47. Krueger RF, Eaton NR. Transdiagnostic factors of mental disorders. World Psychiatry 2015; 14: 27–29.

48. Lee PH, Feng YA, Smoller JW. Pleiotropy and cross-disorder genetics among psychiatric disorders. Biol Psychiatry 2021; 89: 20–31.

49. Grotzinger AD. Shared genetic architecture across psychiatric disorders. Psychol Med 2021; 51: 2210–2216.

50. Cross-Disorder Group of the Psychiatric Genomics Consortium. Identification of risk loci with shared effects on five major psychiatric disorders: a genome-wide analysis. Lancet 2013; 381: 1371–1379.

51. Adamaszek M et al. Consensus paper: cerebellum and emotion. Cerebellum 2017; 16: 552–576.

52. Andreasen NC, Paradiso S, O’Leary DS. “Cognitive dysmetria” as an integrative theory of schizophrenia: a dysfunction in cortical-subcortical-cerebellar circuitry? Schizophr Bull 1998; 24: 203–218.

53. Kabir ZD et al. Cacna1c in the prefrontal cortex regulates depression-related behaviors via REDD1. Neuropsychopharmacology 2017; 42: 2032–2042.

54. Scott AJ, Webb TL, Martyn-St James M, Rowse G, Weich S. Improving sleep quality leads to better mental health: a meta-analysis of randomised controlled trials. Sleep Med Rev 2021; 60: 101556.

55. Ye YY et al. Internet-based cognitive behavioral therapy for insomnia (ICBT-i) improves comorbid anxiety and depression: a meta-analysis of randomized controlled trials. PLoS One 2015; 10: e0142258.

56. Palagini L et al. CBT-I for prevention and early intervention in mental disturbances: a systematic review and meta-analysis. Sleep Med 2024; 124: 650–658.

57. Major Depressive Disorder Working Group of the Psychiatric Genomics Consortium. Trans-ancestry genome-wide study of depression identifies 697 associations implicating cell types and pharmacotherapies. Cell 2025; 188: 640–652.e9.

58. O’Connell KS et al. Genomics yields biological and phenotypic insights into bipolar disorder. Nature 2025; 639: 968–975.

59. Suzuki K et al. Genetic drivers of heterogeneity in type 2 diabetes pathophysiology. Nature 2024; 627: 347–357.

60. Aragam KG et al. Discovery and systematic characterization of risk variants and genes for coronary artery disease in over a million participants. Nat Genet 2022; 54: 1803– 1815.

61. Schoeler T, Pingault JB, Kutalik Z. The impact of self-report inaccuracy in the UK Biobank and its interplay with selective participation. Nat Hum Behav 2025; 9: 584– 594.

