## Supplementary Information (methods, results, Figures S1-S2) for "Insomnia and the genetic overlap of sleep with depression and anxiety: a genomic structural equation modelling study"

### Supplementary Methods

#### *1. Input summary statistics*

Insomnia (ukb-b-3957, the UK Biobank sleeplessness/insomnia item), sleep duration (ukb-b-4424) and chronotype (ukb-b-4956) were taken from the MRC-IEU OpenGWAS resource [1], where they were analysed with BOLT-LMM. MDD was the meta-analysis of Howard et al. excluding 23andMe (ieu-b-102) [2]. Anxiety disorders were the public release of the PGC-ANX meta-analysis, which excludes the Utah cohort (122 083 cases and 729 602 controls) [3]. Bipolar disorder was ieu-b-5110 [4]; schizophrenia the public release of the primary multi-ancestry meta-analysis of the PGC (ieu-b-5099; per-variant sample size up to 175 799) [5]; type 2 diabetes the European-ancestry meta-analysis of Xue et al. (GCST006867) [6]; BMI the GIANT and UK Biobank meta-analysis [7,8]; and coronary artery disease GCST005195 [9]. Summary statistics were munged in GenomicSEM against the HapMap3 variant list with a minor allele frequency threshold of 0.01.

Post-traumatic stress disorder was not used as an indicator of the internalizing factor. Its diagnostic criteria include sleep disturbance [10], so its inclusion would bring sleep-related variance into the factor whose relationship with sleep is the object of the analysis. A case–control PTSD GWAS [11] was used only in the sensitivity analysis with an eleven-indicator covariance matrix estimated separately.

#### *2. Sample size and prevalence choices*

Effective sample sizes were used for the case–control traits, so the sample prevalence was set at 0.5 when converting heritability to the liability scale. The schizophrenia release carries the total sample size (74 776 cases and 101 023 controls at the largest per-variant N), which corresponds to an effective sample size of 171 883. Rescaling a trait by a constant multiplies its genetic variance by that constant and its genetic covariances by the square root, which a standardized model absorbs. We verified this by refitting Model D after rescaling schizophrenia to its effective sample size and anxiety disorders to a population prevalence of 0.20: $\chi^2$, AIC, the insomnia cross-loading and $\Phi$ were unchanged (Supplementary Table S3).

#### *3. Model specification and admissibility*

Models were fitted with usermodel() in GenomicSEM (estimation = "DWLS", std.lv = TRUE). In lavaan syntax, Model C was

Sleep =~ NA*insomnia + sleep_dur + chronotype; Intern =~ NA*mdd + strom_anx; PsychBP =~ NA*bip + scz; Metab =~ NA*t2d + bmi + cad; with each factor variance fixed at 1.

Model D added Intern =~ insomnia. The domain accounts added Intern =~ sleep_dur, Intern =~ chronotype, Sleep =~ mdd or Sleep =~ strom_anx; the shared-content accounts added insomnia mdd, insomnia strom_anx, or both with a common label (insomnia a*mdd and insomnia a*strom_anx); the comparators added PsychBP =~ insomnia or Metab =~ insomnia. A solution was admissible if every standardized loading was at most 1 in absolute value and every residual variance was positive. We also fitted, from Model C, a residual covariance between each pair of sleep indicators, as a repair that does not involve the internalizing domain. Parameter estimates of every model are given in Supplementary Table S23.

The root mean square error of approximation rescales $\chi^2$ by a sample size, and an LDSC sampling covariance matrix has no single sample size because each of its elements rests on a different effective N; we therefore used CFI, SRMR and AIC.

#### *4. Monte Carlo over the LDSC sampling distribution*

Five hundred genetic covariance matrices were drawn from a multivariate normal distribution centred on the estimated matrix, with the LDSC sampling covariance V (seed 20260827). Model D, the domain account with sleep duration, the shared-content account with equal residual covariances, the two comparator placements, the chronotype extension of Model D and Model C were refitted to every draw. The sign of a factor is not identified: in 58 draws for Model D and 56 for its chronotype extension, the Sleep factor emerged reflected, with all its loadings and correlations reversed in sign. Reflection leaves fit and every share unchanged, and these draws were re-aligned to the orientation of the observed solution rather than discarded. All 500 draws entered the summaries (Supplementary Table S5).

#### *5. Decomposition of the cross-domain covariance*

For a sleep indicator *s* and an internalizing indicator *i*, the model-implied genetic covariance $T_{si}$ is the sum over pairs of factors *F*, *G* of $\lambda_{Fs}\ \Phi_{FG}\ \lambda_{Gi}$, plus any residual covariance between *s* and *i*. Terms with *F* = Sleep and *G* = Internalizing form the factor-correlation part; terms in which the sleep indicator loads on a factor other than Sleep form the cross-loading part, attributed to insomnia when *s* is insomnia. The magnitude-weighted insomnia share is Σ sign($T_{si}$) $I_{si}$ / Σ |$T_{si}$| and the net signed share Σ $I_{si}$ / Σ $T_{si}$, both over the six pairs (Supplementary Table S11).

#### *6. Multivariate GWAS and heterogeneity*

The multivariate GWAS used userGWAS under Model D, with the model string read from the file written by the confirmatory analysis so that the fitted and tested models could not diverge. Variants on the sex chromosomes and in the extended major histocompatibility complex were excluded. Genome-wide significance was $P < 5 \times 10^{-8}$; $P < 1.25 \times 10^{-8}$ corrects for four factors. The model-level $Q_{SNP}$ is $\chi^2$ of the common-pathway model minus $\chi^2$ of a model in which the variant acts on each of the ten indicators, with 6 df [12]. For the factor-specific Q [13], the comparison model let the variant act directly on the indicators of the factor tested (Sleep: insomnia, sleep duration, chronotype; Internalizing: MDD, anxiety, insomnia; Psychotic-Bipolar: bipolar disorder, schizophrenia; Metabolic: type 2 diabetes, BMI, coronary artery disease) while still acting on the other three factors (2, 2, 1 and 2 df). The MDD and bipolar tests added a single direct path from the variant to MDD or to bipolar disorder (1 df). Inflation of the factor statistics was summarized by $\lambda_{GC}$ and by an LDSC intercept from regressing factor $\chi^2$ on European HapMap3 LD scores, estimated on variants with $\chi^2 \leq 30$; the intercept is identified by the offset of the regression and needs no sample size (Supplementary Table S15).

#### *7. Locus novelty*

Lead positions were lifted from GRCh37 to GRCh38 with the UCSC hg19ToHg38 chain (pyliftover), and the distance to the nearest catalogued association on the same chromosome was computed against the GWAS Catalog (May 2026 release) [14], which reports GRCh38 positions, without trait filtering. Whether each lead rsID itself appears in the Catalog was recorded as well (Supplementary Table S16).

#### *8. FinnGen replication*

FinnGen R12 endpoints were F5_INSOMNIA, F5_DEPRESSIO, F5_ANXIETY, F5_SCHZPHR and T2D [15]. Variants were matched by rsID and alleles harmonized to the factor effect allele. Direction concordance was tested against 50% with a two-sided binomial test, and effect sizes were correlated across matched leads. The orientation of each factor, fixed by its highest-loading indicator, agreed with the empirical orientation for every factor (Supplementary Table S18). For the overlap analysis, the FinnGen endpoints were munged with effective sample sizes from the R12 manifest and entered a fifteen-trait LDSC run with the ten inputs; the cross-trait intercept between two GWAS is non-zero when they share participants.

#### *9. Fine-mapping and colocalization*

For the two strongest leads of each factor, LD between variants within 250 kb was computed in the 1000 Genomes European reference and the consistency of the summary statistics with that LD was assessed with estimate_s_rss (susieR), at a bound of 0.05. Loci within the bound were fine-mapped with susie_rss (L = 10), with credible sets at 95% coverage. For colocalization, full summary statistics for GTEx v8 cerebellar hemisphere, cerebellum and frontal cortex (BA9) were obtained from the eQTL Catalogue [16] (datasets QTD000161, QTD000166 and QTD000176; 175, 209 and 175 donors) for a 600-kb window around each lead (GRCh38) and matched to the GWAS by rsID. coloc.abf [17] was run for every gene with at least 50 shared variants, with sdY = 1 for both traits, the GTEx sample size for expression, the default priors and, as a sensitivity analysis, $p_{12} = 5 \times 10^{-6}$ [18]. SuSiE-based colocalization (coloc.bf_bf) compared the GWAS credible sets with the eQTL Catalogue's in-sample SuSiE fine-mapping of each gene [19]. Fine-mapping requires a sample size, which a factor GWAS lacks; we used 500 199 and repeated the screening and fine-mapping across a 5.4-fold range (150 670 to 810 294): every screening decision was unchanged, and posterior inclusion probabilities changed by less than 0.001 (Supplementary Table S9).

#### *10. Gene-based and tissue analyses*

MAGMA [20] gene-based tests used the snp-wise mean model with GENCODE v19 genes [21], a 10-kb window and within-factor Bonferroni correction across 17 151 genes; the gene-property analysis used GTEx v8 expression across 54 tissues [22], with Bonferroni correction across tissues (Supplementary Table S8).

#### *11. Software and code*

R 4.4.3 with GenomicSEM 0.0.5 (lavaan 0.6.21), susieR 0.14.2, coloc 5.2.3, data.table 1.17.0 and MASS 7.3.64, run in an Apptainer container on the TRUBA cluster; Python 3.11 with pandas, matplotlib and pyliftover for annotation and figures. The scripts that produce every number, table and figure are available from the corresponding author on request. Figures and tables are generated from the result files without hand-entered values.

### Supplementary Results

#### *1. Prespecified sensitivity models*

Unconstrained intercepts cannot be fitted to an LDSC covariance matrix, which has no mean structure. Collapsing Sleep into Internalizing fitted far worse than Model D (CFI = 0.841, SRMR = 0.091, AIC = 1 617 against 388), and a bifactor structure did not yield an admissible solution (largest standardized loading 2.076; a residual variance of −7.23) (Supplementary Table S24).

#### *2. Repairs within the sleep domain*

Residual covariances between pairs of sleep indicators, added to Model C, did not restore admissibility: with insomnia–chronotype or sleep duration–chronotype the insomnia loading on Sleep remained 1.196 and 1.199, and the model with an insomnia–sleep duration residual covariance did not converge (Supplementary Table S13). The inadmissibility of Model C therefore concerns how insomnia relates to the internalizing indicators, not the covariance among the sleep indicators.

#### *3. Admissibility across Monte Carlo draws*

Model D and the domain account were admissible in 98.6% of the 500 draws, the shared-content account in 78.0%, the Psychotic-Bipolar placement in 4.2% and Model C in none; the Metabolic placement was inadmissible in all 464 draws in which it converged (Supplementary Table S5).

#### *4. Fine-mapping*

The four loci within the LD-mismatch bound (s = 0.006–0.022) were fine-mapped; the four outside it (s = 0.100–0.707) were not (Supplementary Table S21; Supplementary Fig. S2a). At the Internalizing locus on chromosome 5 (rs60271) a single credible set placed most of the posterior on the lead variant (PIP = 0.806). At *CACNA1C* (rs4298967) the lead was the top variant (PIP = 0.761; two credible sets). The Sleep locus on chromosome 2 (rs1950160) resolved to rs10490095 (PIP = 0.645; three credible sets), and the Internalizing locus on chromosome 1 (rs3101336) spread over two credible sets (largest PIP 0.371, rs2568958) (Supplementary Fig. S2b).

#### *5. Colocalization*

At the chromosome 1 Internalizing locus, the association colocalized with expression of ENSG00000286863, a long non-coding RNA without a GENCODE v19 name that Ensembl release 111 maps to *LINC02796*, in frontal cortex (PP.H4 = 0.995), cerebellar hemisphere (0.994) and, less clearly, cerebellum (0.674); the result held at $p_{12} = 5 \times 10^{-6}$ (0.991 and 0.988) and in SuSiE-based colocalization (0.992 and 0.990). The lead variant lies upstream of *NEGR1*, whose expression did not colocalize (PP.H4 = 0.23 in frontal cortex, 0.30 in cerebellar hemisphere and 0.07 in cerebellum). At *CACNA1C*, single-variant colocalization favoured distinct causal variants (PP.H3 = 0.83 in cerebellar hemisphere and 0.99 in cerebellum). SuSiE-based colocalization separated the signals: the GWAS signal tagged by rs4298967 colocalized with a *CACNA1C* eQTL signal in cerebellar hemisphere (PP.H4 = 0.991) and cerebellum (0.713), whereas the second GWAS signal and the second eQTL signal did not match (PP.H3 ≥ 0.97). The Sleep locus on chromosome 2 and the Internalizing locus on chromosome 5 showed no colocalization (PP.H4 < 0.5 for every gene) (Supplementary Table S22; Supplementary Fig. S2c).

#### *6. Gene-based and tissue enrichment*

After within-factor Bonferroni correction, MAGMA identified 110 genes for Sleep, 230 for Internalizing, 510 for Psychotic-Bipolar and 1 505 for Metabolic. All four factors were enriched in brain tissues (13 or 14 of 54 tissues per factor): frontal cortex ranked first for Sleep ($P = 6.2 \times 10^{-14}$) and Internalizing ($P = 9.1 \times 10^{-13}$), and cerebellar hemisphere for Psychotic-Bipolar ($P = 4.3 \times 10^{-33}$) and Metabolic ($P = 6.2 \times 10^{-19}$) (Supplementary Table S8). Because the same tissues lead for all four factors, these enrichments are not specific to any one of them.

### Supplementary Figures

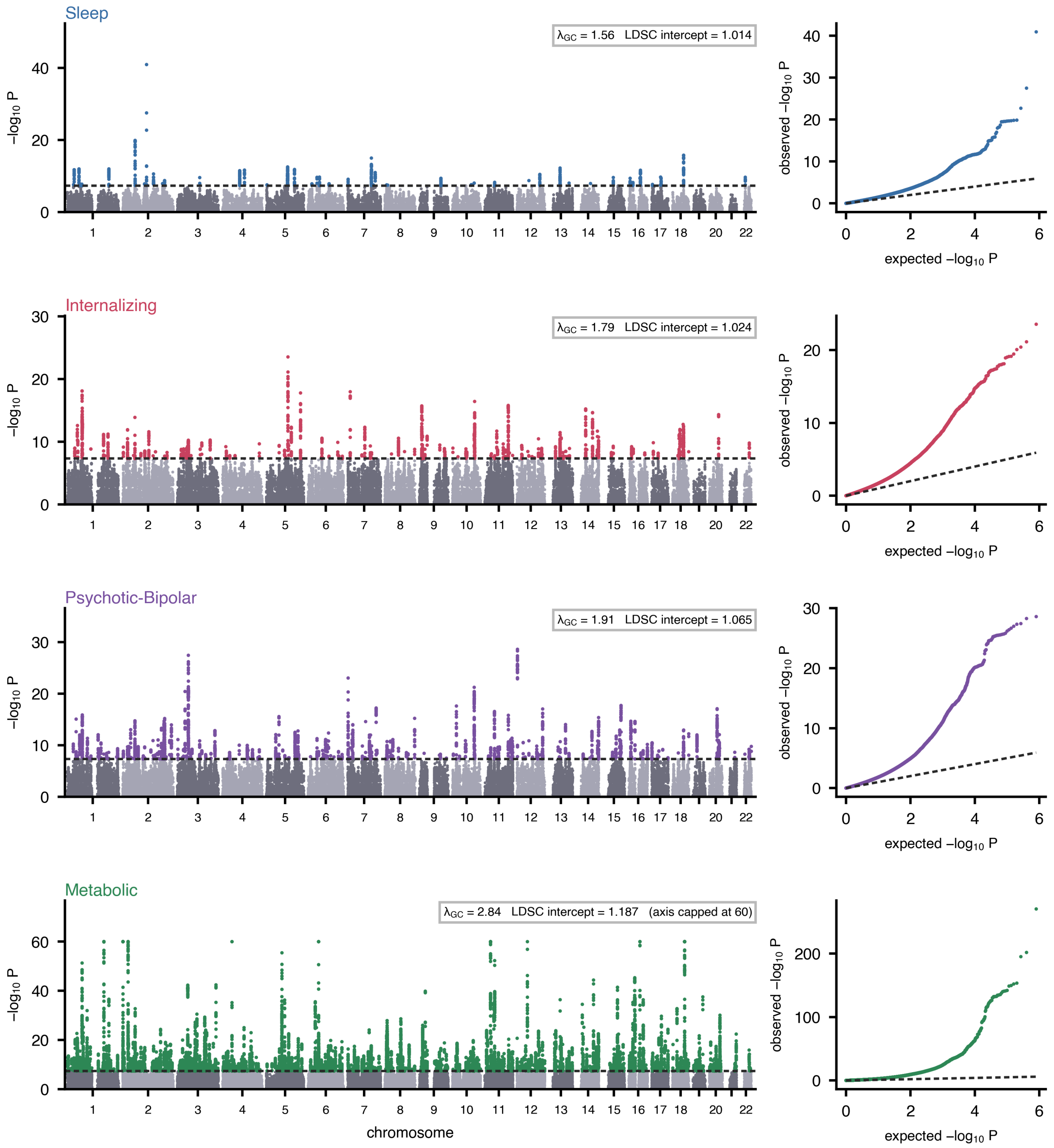


**Supplementary Figure S1. Genome-wide results for the four factors.** Manhattan plots (left; the Metabolic axis is capped at $-\log_{10}$ P = 60) and quantile–quantile plots (right), with $\lambda_{GC}$ and the LDSC intercept of each factor. The dashed line marks $P = 5 \times 10^{-8}$.

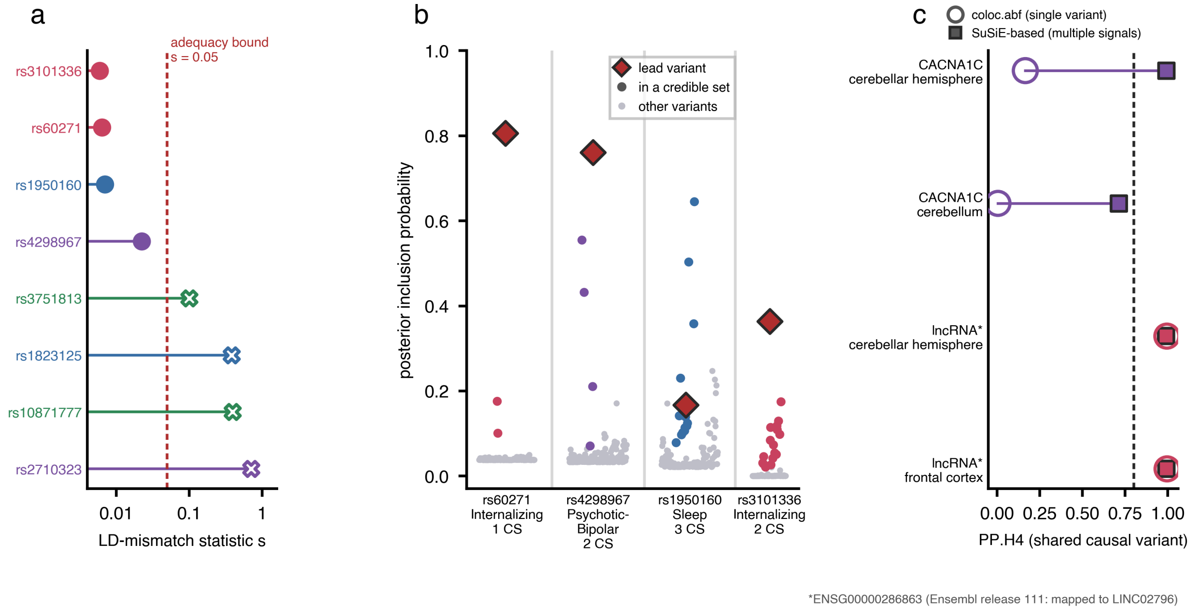


**Supplementary Figure S2. Locus follow-up.** (**a**) LD-mismatch statistic for the two strongest leads of each factor against the bound of 0.05; crosses mark loci outside it. (**b**) Posterior inclusion probabilities in the four fine-mapped windows, with the lead variant marked and credible-set members coloured. (**c**) Posterior probability of a shared causal variant (PP.H4) from single-variant colocalization (open circles) and SuSiE-based colocalization (squares) for *CACNA1C* and for ENSG00000286863, whose Ensembl release 111 name is *LINC02796*.

### Supplementary references


1. Elsworth B et al. The MRC IEU OpenGWAS data infrastructure. Preprint at bioRxiv https://doi.org/10.1101/2020.08.10.244293 (2020).

2. Howard DM et al. Genome-wide meta-analysis of depression identifies 102 independent variants and highlights the importance of the prefrontal brain regions. Nat Neurosci 2019; 22: 343–352.

3. Strom NI et al. Genome-wide association study of major anxiety disorders in 122,341 European-ancestry cases identifies 58 loci and highlights GABAergic signaling. Nat Genet 2026; 58: 275–288.

4. Mullins N et al. Genome-wide association study of more than 40,000 bipolar disorder cases provides new insights into the underlying biology. Nat Genet 2021; 53: 817–829.

5. Trubetskoy V et al. Mapping genomic loci implicates genes and synaptic biology in schizophrenia. Nature 2022; 604: 502–508.

6. Xue A et al. Genome-wide association analyses identify 143 risk variants and putative regulatory mechanisms for type 2 diabetes. Nat Commun 2018; 9: 2941.
7. Pulit SL et al. Meta-analysis of genome-wide association studies for body fat distribution in 694 649 individuals of European ancestry. Hum Mol Genet 2019; 28: 166–174.
8. Locke AE et al. Genetic studies of body mass index yield new insights for obesity biology. Nature 2015; 518: 197–206.
9. van der Harst P, Verweij N. Identification of 64 novel genetic loci provides an expanded view on the genetic architecture of coronary artery disease. Circ Res 2018; 122: 433–443.
10. American Psychiatric Association. Diagnostic and Statistical Manual of Mental Disorders, 5th edn (American Psychiatric Publishing, Arlington, VA, 2013).
11. Nievergelt CM et al. Genome-wide association analyses identify 95 risk loci and provide insights into the neurobiology of post-traumatic stress disorder. Nat Genet 2024; 56: 792–808.
12. Grotzinger AD et al. Genomic structural equation modelling provides insights into the multivariate genetic architecture of complex traits. Nat Hum Behav 2019; 3: 513–525.
13. Grotzinger AD et al. Genetic architecture of 11 major psychiatric disorders at biobehavioral, functional genomic and molecular genetic levels of analysis. Nat Genet 2022; 54: 548–559.
14. Sollis E et al. The NHGRI-EBI GWAS Catalog: knowledgebase and deposition resource. Nucleic Acids Res 2023; 51: D977–D985.
15. Kurki MI et al. FinnGen provides genetic insights from a well-phenotyped isolated population. Nature 2023; 613: 508–518.
16. Kerimov N et al. A compendium of uniformly processed human gene expression and splicing quantitative trait loci. Nat Genet 2021; 53: 1290–1299.
17. Giambartolomei C et al. Bayesian test for colocalisation between pairs of genetic association studies using summary statistics. PLoS Genet 2014; 10: e1004383.
18. Wallace C. Eliciting priors and relaxing the single causal variant assumption in colocalisation analyses. PLoS Genet 2020; 16: e1008720.

19. Wallace C. A more accurate method for colocalisation analysis allowing for multiple causal variants. PLoS Genet 2021; 17: e1009440.

20. de Leeuw CA, Mooij JM, Heskes T, Posthuma D. MAGMA: generalized gene-set analysis of GWAS data. PLoS Comput Biol 2015; 11: e1004219.

21. Frankish A et al. GENCODE reference annotation for the human and mouse genomes. Nucleic Acids Res 2019; 47: D766–D773.

22. GTEx Consortium. The GTEx Consortium atlas of genetic regulatory effects across human tissues. Science 2020; 369: 1318–1330.

**Supplementary Tables**

The Supplementary Tables are provided as a single Excel workbook (Supplementary_Tables.xlsx), one sheet per table.

**Supplementary Table S1. SNP heritability and single-trait LDSC statistics of the ten input GWAS.** Population prevalence is the value used for liability-scale conversion of case–control traits (sample prevalence 0.5). The N entering LDSC is the per-variant sample size of the munged file; for schizophrenia it is the total rather than the effective sample size (Supplementary Methods). The h2 Z is computed by LDSC from unrounded estimates.

**Supplementary Table S2. LDSC genetic correlations among the ten input GWAS.** Two-sided P from the Z statistic.

**Supplementary Table S3. Invariance of the standardized model to the scale of single indicators.**

**Supplementary Table S4. Fit of Models A–D.** Admissible: all standardized loadings at most 1 in absolute value and all residual variances positive.

**Supplementary Table S5. Parametric Monte Carlo over the LDSC sampling distribution.** Mirror solutions, in which the Sleep factor emerged with all its loadings and correlations reversed in sign, were re-aligned rather than discarded (58 draws for Model D and 56 for its chronotype extension); fit and shares are unaffected by the reflection.

**Supplementary Table S6. Model D with residual variances of the Internalizing indicators fixed.**

**Supplementary Table S7. Leave-one-indicator-out refits of Model D.**

**Supplementary Table S8. Gene-based and tissue enrichment analyses.**

**Supplementary Table S9. Sensitivity of fine-mapping to the assumed factor sample size.**

**Supplementary Table S10. Full parameter estimates of Model D.** Diagonally weighted least squares; factor variances fixed at 1.

**Supplementary Table S11. Decomposition of the six model-implied Sleep × Internalizing covariances.**

**Supplementary Table S12. Post-traumatic stress disorder as a third Internalizing indicator.** Eleven-indicator LDSC covariance matrix estimated separately; PTSD entered no other analysis.

**Supplementary Table S13. Every one-parameter link added to Model C, its comparators and its two-link extensions.** Each row adds the listed parameter(s) to Model C. Weighted insomnia share: magnitude-weighted share of the six model-implied Sleep × Internalizing covariances carried by the insomnia cross-loading.

**Supplementary Table S14. Factor lead variants.**

**Supplementary Table S15. Inflation and LDSC calibration of the factor GWAS.**

**Supplementary Table S16. Locus novelty against the GWAS Catalog on a single genome build.**

**Supplementary Table S17. Heterogeneity of factor lead variants against the model.** Lead-set Bonferroni: 0.05 divided by the number of leads of the factor. Genome-wide Bonferroni: 0.05/810 294.

**Supplementary Table S18. Replication of factor lead variants in FinnGen R12.**

**Supplementary Table S19. Replication of indicator lead variants in FinnGen R12.**

**Supplementary Table S20. Sample overlap between the input GWAS and FinnGen R12 endpoints.**

**Supplementary Table S21. LD-mismatch screening and fine-mapping.**

**Supplementary Table S22. Colocalization of fine-mapped loci with brain expression.** Tissues: GTEx v8 cerebellar hemisphere, cerebellum and frontal cortex (BA9).

**Supplementary Table S23. Parameter estimates of every model in the rival-model table.**

**Supplementary Table S24. Prespecified sensitivity models.**
